# Class I HLA allele predicted antigenic coverages for Spike and Nucleocapsid proteins are associated with deaths related to COVID-19

**DOI:** 10.1101/2020.06.03.20121301

**Authors:** Marco Antônio M. Pretti, Rômulo G. Galvani, Gustavo Fioravanti Vieira, Adriana Bonomo, Martín H. Bonamino, Mariana Boroni

## Abstract

The world is dealing with one of the worst pandemics ever. SARS-CoV-2 is the etiological agent of COVID-19 that has already spread to more than 200 countries. However, infectivity, severity, and mortality rates do not affect all countries equally. Here we consider 140 HLA alleles and extensively investigate the landscape of 3,723 potential HLA-I A and B restricted SARS-CoV-2-derived antigens and how 37 countries in the world are predicted to respond to those peptides considering their HLA-I distribution frequencies. The clustering of HLA-A and HLA-B allele frequencies partially separates most countries with the lowest number of deaths per million inhabitants from the other countries. We further correlated the patterns of *in silico* predicted population coverage and epidemiological data. The number of deaths per million inhabitants correlates to the predicted antigen coverage of S and N derived peptides and its module is influenced if a given set of frequent or rare HLA alleles are analyzed in a given population. Moreover, we highlighted a potential risk group carrying HLAs associated with an elevated number of deaths per million inhabitants. In addition, we identified 3 potential antigens bearing at least one amino acid of the 4-length insertion that differentiates SARS-CoV-2 from previous coronavirus strains. We believe these data can contribute to the search for peptides with the potential to be used in vaccine strategies considering the role of herd immunity to hamper the spread of the disease. Importantly, to the best of our knowledge, this work is the first to use a populational approach in association with COVID-19 outcome.

**Importance:** The emergence of COVID-19 outbreak caused by a novel coronavirus opens up an imminent need to better understand how our immune system responds to this new virus and to develop ways to control its spread. Our results suggest why some countries show a higher number of deaths due to COVID-19 while other countries do not. SARS-CoV-2 expresses 10 proteins that could be used as targets for vaccine development. By using a comprehensive bioinformatic screening of potential epitopes derived from the SARS-CoV-2 sequences, we identified potential antigens for 148 HLA-I alleles distributed world-wide. These peptides are likely to have a high affinity for HLA class I molecules and may induce critical immune responses. Our results suggest that different coverages for S and N derived peptides is associated with deaths related to COVID-19 in distinct populations. Of note, frequent and rare HLA alleles influence the effects we observed. We explored these associations regarding potential antigens derived from each viral protein to enumerate a set of protective HLA alleles. Moreover, we explored the novel insertion in the SARS-CoV-2 protein S genome to map 3 potential antigens bearing this new region, and a set of peptides presented by those protective HLA alleles, of interest for vaccine strategies. Finally, we propose that vaccine development strategies should consider the inverse relationships of proteins S and N in view of the associations with the number of deaths.

## Introduction

Since December 2019, the world has been facing one of the worst pandemics of all times caused by a novel *Betacoronavirus* (Severe Acute Respiratory Syndrome Coronavirus 2 - SARS-CoV-2). Until this day, more than 20 million cases and 732 thousand deaths were confirmed. However, the infection does not strike all nations equally when considering multiple epidemiological parameters (1). The viral genome of this etiological agent consists of structural protein genes such as Spike (S), Envelope (E), Membrane (M), and Nucleocapsid (N) and non-structural protein genes (e.g., ORF1ab, ORF3a, ORF6) (2). Their functions have been extensively reviewed elsewhere (2,3). Once in the human body, SARS-CoV-2 infect cells using the surface molecules ACE2 and TMPRSS2 highly expressed in the lungs and respiratory tract. The infection is recognized by macrophages and alveolar epithelial cells that initiate a pro-inflammatory response, which may trigger an acute respiratory distress syndrome among other symptoms (4–7). The SARS-CoV-2 viral proteins have been shown to elicit both cellular immunity and humoral response (8). One of the most promising viral proteins for vaccine development is the S protein due to its accessibility to antibodies and pivotal role in the infection (9). Nevertheless, antibody response appears first against the N protein, the most abundant protein in coronavirus, and a few days later for S protein (8,9). The ability to trigger an adaptive immune response greatly relies on the ability to present antigens through the class I and II Histocompatibility Leukocyte Antigen (HLA) molecules. Peptides derived from different SARS-CoV-2 proteins are recognized by CD4 and CD8 T cells from COVID-19 convalescent patients (10). The allelic distribution of the largely polymorphic HLA genes across different countries may affect the appropriate antigen presentation of SARS-CoV-2-derived peptides among distinct populations (11). This could be associated with specific profiles of disease susceptibility and severity (1). Several studies predicting HLA-restricted peptides derived from SARS-CoV-2 are already available (12,13). Therefore, these characteristics must be taken into account when considering epitope-based vaccines since target epitopes must not only be able to bind to the HLA molecule but also prove to be immunogenic to promote a functional response (14). Populational coverage studies considering these features may help narrow the number of candidate peptides to be considered for vaccination strategies (15). However, epitope coverage studies are not sufficient to predict if the immune response would be able to resolve the infection and provide immunity to individuals and/or if this immunization occurs at the expense of exacerbated immune responses. In the context of the SARS-CoV-2 pandemic, the susceptibility to severe cases and deaths may be related to the over or under presentation of SARS-CoV-2-derived peptides by HLA molecules as well as to individual characteristics driving inflammation (3). This is not necessarily restricted to HLA allele distribution among populations once the same peptide can be presented by more than one HLA allele. With that in mind, we evaluated how different populations potentially present HLA class I restricted SARS-CoV-2 peptides and their association with the local epidemic progression features across different countries. This strategy led us to a set of peptides derived from virus S and N proteins, which showed distinct correlations with disease outcomes. While S may have a protective role as they are potentially presented in HLA alleles with high frequency in countries with less number of deaths per million inhabitants, N has a direct correlation with the unfavorable endpoint. Importantly, the patterns observed are dependent on frequent HLA alleles, and opposite correlations are found when evaluating only rare alleles. These results may prove critical in guiding efforts towards the development of vaccine candidates, as well as determine high-risk groups based on HLA alleles.

## Results

### HLA-I alleles partially discriminate different outcomes among countries

To assess the landscape of peptides potentially presented by HLA-I alleles occurring in the global population and how this diversity is associated with the disease outcome, we selected a list of countries hit by the pandemic with known HLAs genetic frequencies. Only countries with more than 1,000 total cases of Coronavirus disease (COVID-19) on 17/05/2020 according to the Worldometer database (16) register were considered, reaching a total of 50 different countries. Two field resolution HLA-A and HLA-B alleles for the aforementioned populations available in the Allele Frequency Net Database (17) were accessed. For each population, a different number of alleles was retrieved, considering the allele in decreasing order of their frequencies to reach a cumulative allelic frequency (AF) as close to 0.75 as possible. Populations whose HLA allele frequencies information did not reach the threshold or having less than 50 individuals in the HLA cohort were removed, totalizing 37 countries in our study (Supplementary File 1, Figure S1). The median cumulative AF varied among populations with a median of 0.752 for HLA-A (minimum of 0.721 for AUT and maximum of 0.781 for GRE) and 0.739 for HLA-B (minimum of 0.714 for SWE and maximum of 0.750 for MEX) and the number of HLA-I alleles ranged from 4 to 14 for HLA-A (median=7) and from 6 to 26 for HLA-B (median=13, Table S1).

To investigate whether different HLA-I alleles among populations could be associated with epidemiologic features (retrieved from the Worldometer database, see methods), we sorted countries in ascending order regarding these data (Supplementary File 1). The USA was the country with the highest number of cases, followed by RUS, ESP, GBR, BRA, and ITA, while the countries with the lowest number of cases were SEN, CZE, TUN, KOR, and BUL (Figure S2A). However, when we evaluate cases per million inhabitants, IRL was the leading country in the rank, followed by SGP, USA, ITA, SWE, and POR while BRA, THA, CHN, INA, TUN, and JPN were the countries with the lowest ratios (Figure S2B). The USA also led the rank in the number of COVID-19 deaths, followed by GBR, ITA, FRA, BRA, and DEU while the countries with the lowest number of deaths were OMA, SGP, SEN, GHA, and TUN (Figure S2C). When analyzing the number of deaths per million inhabitants, ESP was the country with the highest ratio followed by ITA, GBR, FRA, SWE, and HOL while THA, GHA, SEN, CHN, MAL, and INA were the countries with the lowest ratios (Figure S2D). The USA also had the highest number of serious cases followed by BRA, FRA, GBR, DEU, and ESP, while the countries with the lowest number of serious cases were MAR, UAE, TUN, GHA, SEN, and ARM (Figure S2E). On the other hand, the USA also led in the number of recovered, followed by ESP, ITA, BRA, CHN, and RUS while BUL, TUN, SGP, SEN, ROM, and GRE were the countries with the lowest numbers of recovered (Figure S2F).

We chose to use death per million inhabitants as the main epidemiological metric as it is the one that varies least depending on the testing capacity or policy adopted by the country studied (18). To evaluate if the distribution of HLA allele frequencies among populations could explain death per million inhabitants, we performed an unsupervised hierarchical clustering. We used HLA-A and HLA-B frequencies for each particular country, that were divided into quartiles according to the epidemiological parameter (Figure 1). We observed, as expected, that geographic location strongly influences the clustering since cluster 1 is composed exclusively by African countries (pink, RSA, GHA and SEN), corresponding to 60% of all African countries analyzed, with a Jaccard similarity coefficient (JSC) of 0.91, while Cluster 2, also homogeneous regarding the geographic composition, is composed only of Asian countries (yellow, INA, MAL, THA, CHN, and SGP) corresponding to 41% of all Asian countries evaluated in this study, with a JSC of 0.93 (Figure 1). However, we observed that all countries comprising clusters 1 and 2 belong to the first quartile (Q1, green, less number of deaths per million). Therefore, although the HLA frequency clustering recaps the country’s continental distribution, it also appears to separate the first quartile apart from the remaining groups. The most prevalent HLA-I alleles in Q1 are HLA-A*24:02, HLA-A*11:01, HLA-B*40:01, and HLA-A*23:01 while in Q4 (red, more number of deaths per million) are HLA-A*02:01, HLA-A*01:01, HLA-A*03:01, and HLA-B*07:02. Using a Principal Component Analysis (PCA), the same result was observed: despite the clustering by continents, countries belonging to the first quartile (green) are separated from those of the other quartiles (Figure S3). This result highlights the contribution of HLA alleles to segregate distinct responses to the COVID-19 infection among countries. We reasoned that describing the binding spectra of each allele for peptides related to SARS-CoV-2 could further contribute to a better understanding of the epidemiologic data. Therefore, we next sought to perform a thorough characterization of SARS-CoV-2 derived peptide binding prediction to HLA-I alleles.

**Figure 1.**
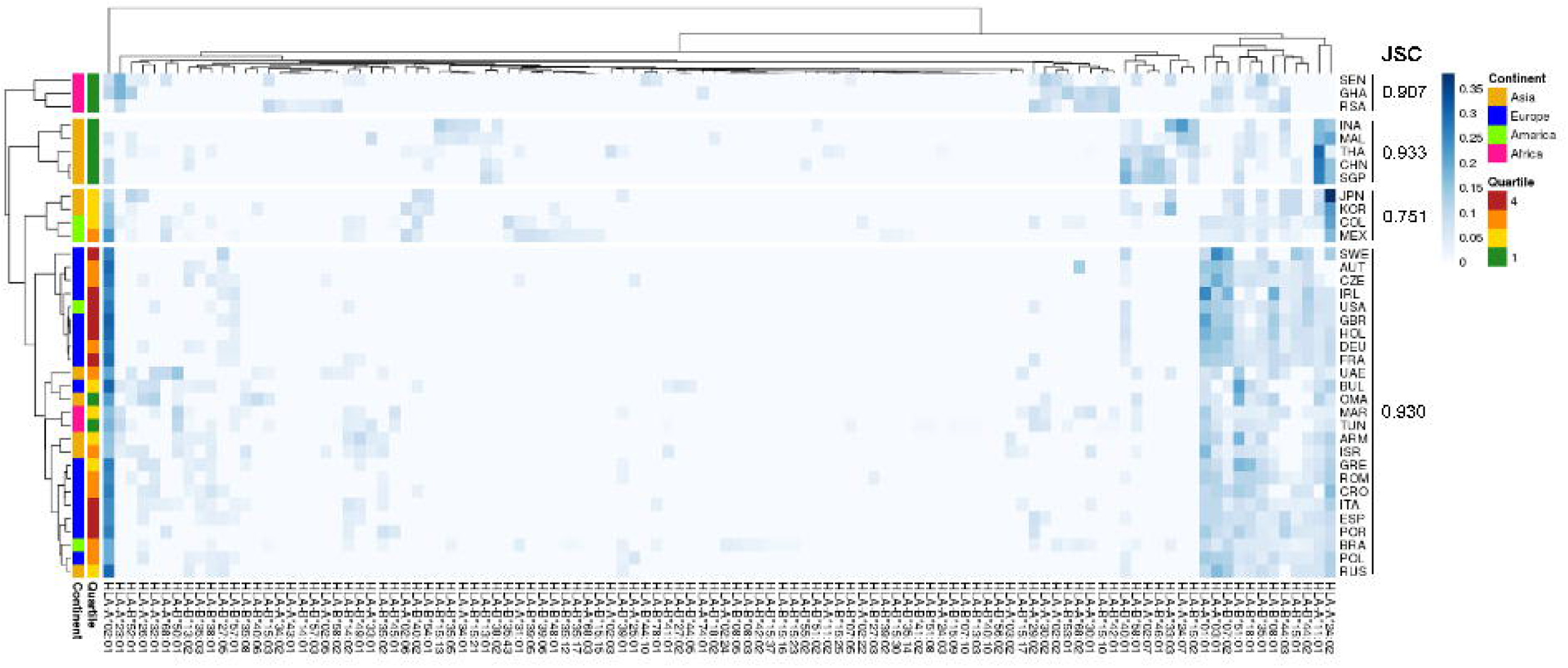
Unsupervised two-dimensional cluster analysis of 37 countries by HLA-A and HLA-B Allele frequencies. Each row represents a population, the first column represents the deaths per million inhabitants quartiles: Q1 = green (lower than 5); Q2 = yellow (5 to 20); Q3 = orange (21 to 85); Q4 = red (greater than 85). The other columns represent HLA alleles, which frequencies values are represented by the blue scale color bar. The complete method and 1-Pearson correlation coefficient as distance were used to cluster rows and columns.

### HLA-I binding of SARS-CoV-2-derived peptides among different populations

In order to study the binding of SARS-CoV-2 peptides to different HLA alleles, we predicted the affinities of HLA alleles (33 HLA-A and 73 HLA-B, Supplementary File 2) for each potential SARS-CoV-2 derived peptide using the netMHCpan4 tool (19). Only peptides that met the criteria of scoring high probabilities of being transported by TAP, and processed by the proteasome (predicted both by netCTLpan (20)) were considered as the final dataset (Fig S4A-B). The 38,464 predicted 8-11 mer unique peptides were derived from the complete genome of the SARS-CoV-2. Of those, 18,799 unique peptides passed the TAP transport score, 12,230 the proteasome cutting, and 10,909 both filters (Fig S4C). Peptides were categorized as Strong Binders (SB, %Rank < 0.5), Weak Binders (WB, 0.5 ≤ %Rank < 2), and Non-binder (NB, %Rank ≥ 2) based on the %Rank definition by the developers, which accounts for the probability of a peptide to bind the HLA given a pool of natural ligands. We confirmed that the number of SB peptides increases with the number of HLA-I alleles accordingly for a given population by performing a Pearson correlation between these two variables (R=0.95, p-value < 0.01, Figure S4D). Even considering that this correlation could be expected, it is important to emphasize that the number of SB peptides restricted to a given HLA allele varies greatly and shall affect a given population antigen coverage.

When evaluating the peptides according to its predicted affinity to HLA alleles, a crescent value is observed with the decrease in the qualitative classifications according to the program, i.e., lower nM affinity for peptides classified as SB (median of 181.1 nM), moderate for WB (median of 1,571.4 nM) and higher for NB (median of 29,660.2 nM, Figure S5A). In order to define which group of peptides we should work with, we compared the predicted *in silico* affinities with affinities of viral-derived peptides obtained by experimental assays. The aforementioned data was retrieved from the Immune Epitope Database (IEDB). A total of 49,207 MHC Ligand assays were retrieved following the selection criteria: 38,558 for HLA-A and 10,649 for HLA-B, classified by the IEDB according to the results of *in vitro* assays readout into Positive-High (Pos-H, 17,353), Positive-Intermediate (Pos-I, 14,927) or Positive-Low (Pos-L, 16,927). The IEDB classifications take into account the information provided in deposited papers: peptides categorized into Pos-L/-I/-H have been grouped according to an IC50 threshold. A similar pattern was observed for the experimental affinities which were lower for Pos-H peptides, a median of 10.1 nM, and higher for Pos-L peptides, a median of 1,430 nM (Wilcox, p < 0.0001, Figure S5B). Moreover, we also predicted the binding affinities of 18,522 unique peptide/HLA pairs retrieved from the IEDB whose capacities to elicit a TCR response were tested through 34,872 T cell assays. They are qualitatively classified into Positive (Pos, 2,705) or Negative (Neg, 15,817) taking into account *in vitro* assays such as ELISPOT and chromium release. A significant difference in the affinities was observed when comparing Pos peptides affinities assays (median of 49.1 nM) with Neg (median of 188.0 nM) ones (Wilcox, p < 0.0001, Figure S5C). It is important to mention that a difference in affinity (Wilcoxon, p < 0.0001) was observed for the predicted data (Figure S5D) but no overall difference in binding affinities was observed for IEDB HLA-A and B alleles (Figure S5E). Half of the HLA alleles assessed in the *in vitro* binding assays selected from the IEDB are shared by the HLAs used for predictions in our analysis. In summary, the predicted affinities of peptides classified into strong, weak or no binder are comparable to the experimental affinities from ligand assays at the IEDB, showing that SB peptides have predicted affinities equivalent to the vast majority of peptides that were proven to bind the HLA experimentally (Pos-H and Pos-I - Figures S5A-B), potentially eliciting an immune response. Therefore, we decided to work only with the SB peptides in the downstream analyses, totalizing 3,723 peptides that passed both TAP and proteasome filtering (Fig S4C).

Considering all SB peptides predicted to bind to the 106 analyzed alleles, none of them matched with 100% identity the sequences found in the human proteome using BLAST search. Different HLA alleles can bind to different peptide repertoires, which is defined by the amino acids that compose the cleft. Alleles that bind to a large repertoire of peptides are called generalists and those that bind to a smaller repertoire of peptides can be called specialists (21). The HLA-A alleles with the capacity to present the higher number of SB peptides are HLA-A*43:01 (302 unique peptides), HLA-A*01:01 (300 unique peptides), and HLA-A*26:01 (296 unique peptides) in contrast to HLA-A*31:01 (84 unique peptides), which is predicted to present the lowest number of SB peptides (Figure 2A). Searching for specific virus proteins and regions within the virus ligandome, we analyzed the set of potential HLA-restricted peptides per population and open reading frames (ORF). The number of SB peptides potentially presented by each allele varies greatly depending on the virus ORF considered, and it is proportional to ORF sizes (Figures 2B, Table S2). Some SB peptides were predicted to be presented by multiple HLA alleles and, as so, evidentiate the discrepancy between the number of peptide:HLA pairs and the number of unique SB peptides derived from viral ORFs (Table S2). For instance, HLA-A*01:01 is predicted to present the highest number of SB peptides (43 unique peptides) derived from protein S (1,274 amino acids - aa) and a moderate number of SB peptides (10 unique peptides) derived from the protein N (420 aa, Figure 2B). Two of the top five most promiscuous (generalist) HLA-A regarding SARS-CoV-2 are country-specific (HLA-A*43:01 from RSA and HLA-A*01:03 from UAE). The most generalist HLA-A (HLA-A*01:01) is found in low frequency in countries that comprise the first quartile (median AF = 0.000, mean=0.0252) and high allele frequency in countries from the fourth quartile (median AF = 0.130, mean=0.148). Regarding the more SARS-CoV-2 fastidious (specialists) HLA-A, HLA-A*33:03 is present in countries that comprise the first quartile (median AF = 0.021, mean=0.0468), but is absent in countries belonging to the fourth quartile. The globally common HLA-A*02:01 is predicted to bind a low number of SB peptides from protein S and N when compared to the other alleles. On the other hand, it is one of the alleles with the higher number of SB peptides from the ORF1ab region, thus being a specialist allele for this region. In contrast, the HLA-A*11:01, A*23:01 and A*24:02 alleles (more prevalent in the countries belonging to the first quartile) seem to act as specialized alleles for the S regions, having the potential to present a greater number of peptides from this region than from the N region (Figure 2B). As for HLA-A, the predicted binding spectrum for HLA-B varies among alleles in which HLA-B*15:21, HLA-B*15:15 and HLA-B*35:43 possess the higher number of SB peptides (286, 269 and 268 unique peptides, respectively), and HLA-B*45:01 (45 unique peptides) the lowest (Figure 3A). The globally commons HLA-B*07:02 and HLA-B*44:02 are predicted to bind low numbers of SB peptides from protein S and N. In contrast, the HLA-B*15:02 (only found in countries belonging to the first quartile) is predicted to bind high numbers of SB proteins from the S region while having the potential to present low numbers of peptides from N protein. (Figure 3B).

**Figure 2.**
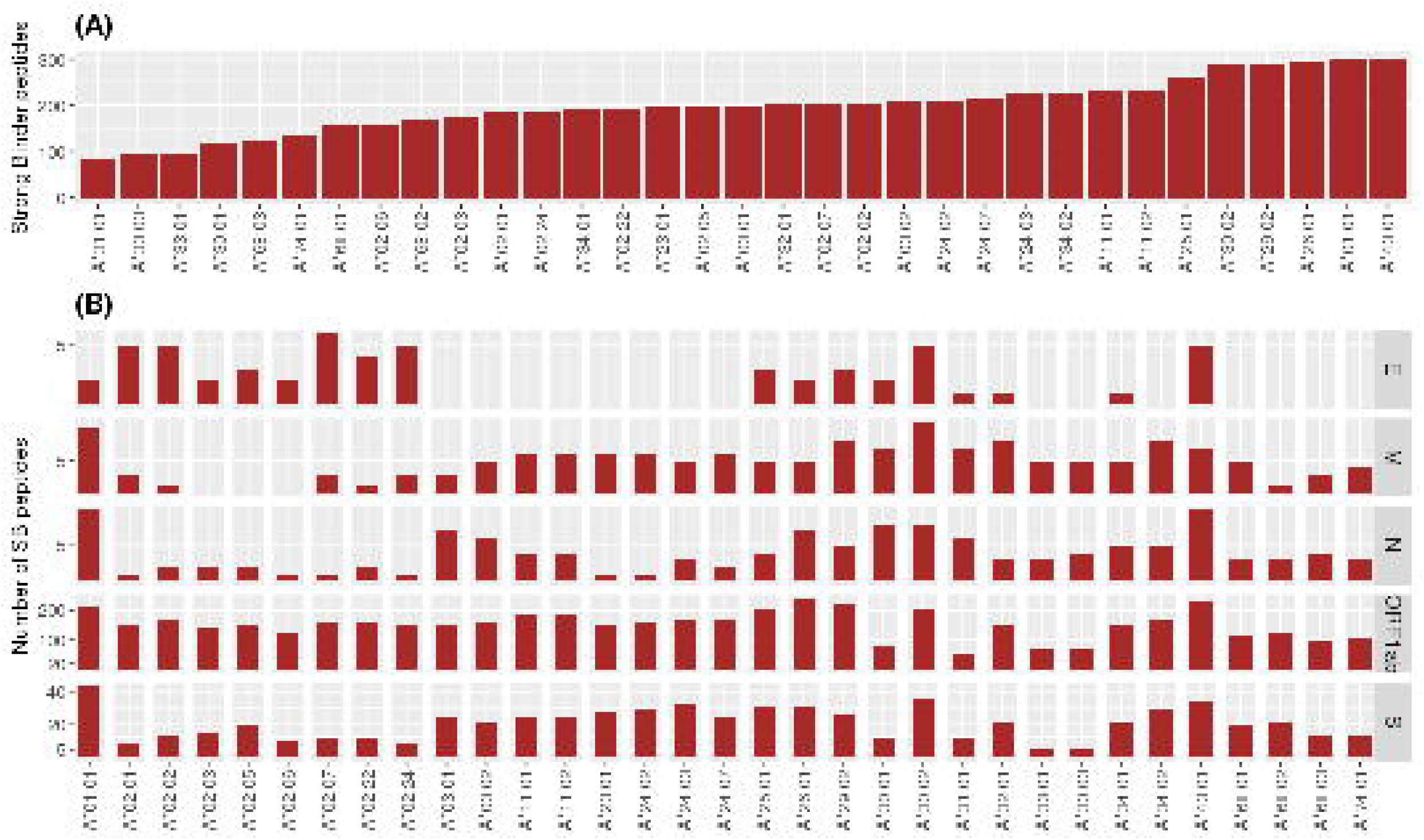
Number of unique Strong Binder peptides for the 33 analyzed HLA-A alleles. **(A)** Global view of the predicted binding spectra for HLA-A considering the entire SARS-CoV-2 genome or **(B)** depicted by ORF1ab, Envelope (E), Membrane(M), Nucleocapsid (N) or Spike (S) proteins. Strong Binder peptides were selected using a %Rank cut-off of 0.5%, which is based on the likelihood of this peptide being presented when compared to a pool of natural ligands. HLA binding predictions performed by netMHCpan were crossed with TAP and proteasome predictions performed by netCTLpan in order to get a more reliable set of peptides. Importantly, the same peptide can be predicted to be presented by more than one HLA.

**Figure 3.**
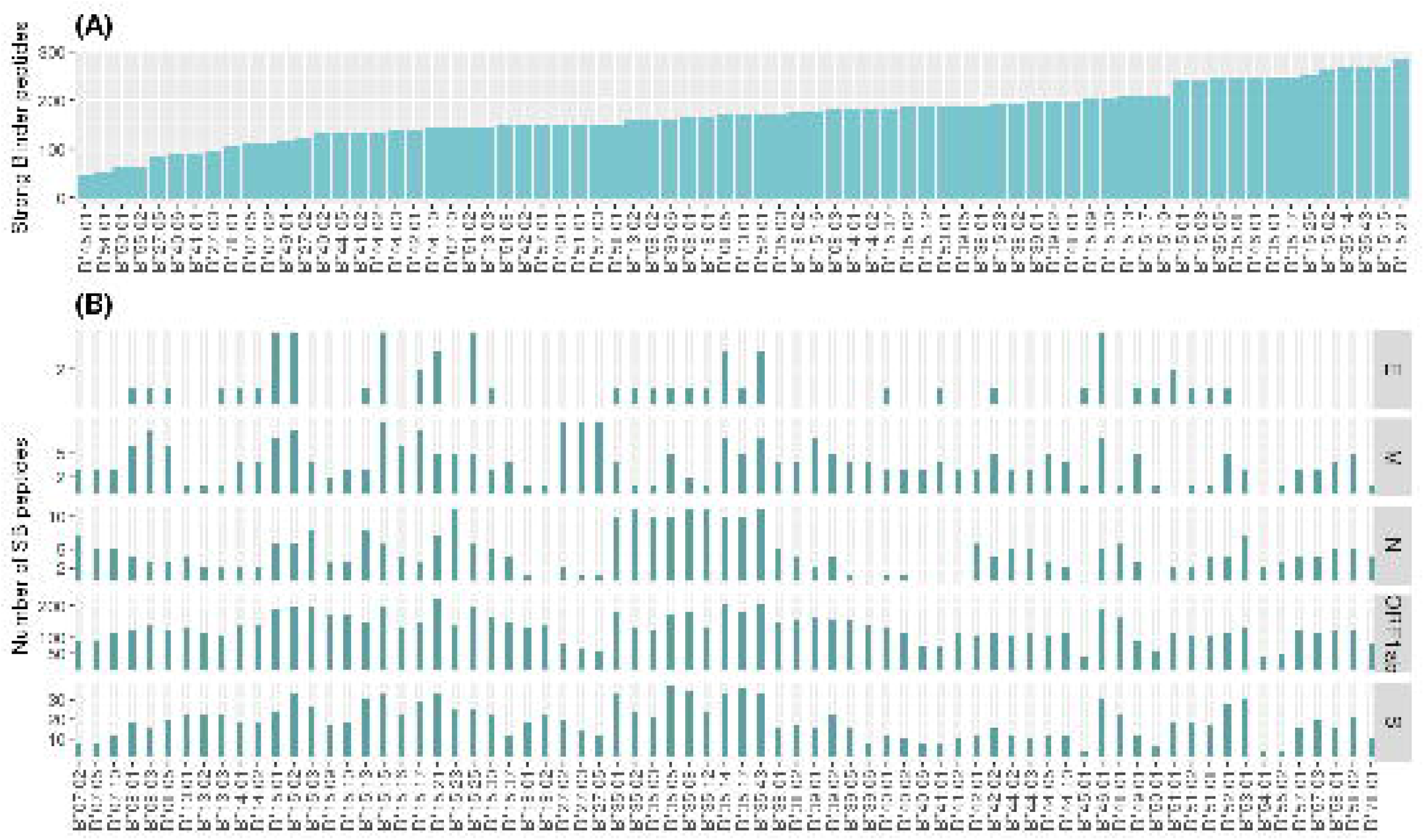
Number of unique Strong Binder peptides for the 76 analyzed HLA-B alleles. **(A)** Global view of the predicted binding spectra for HLA-A considering the entire SARS-CoV-2 genome or **(B)** depicted by ORF1ab, Envelope (E), Membrane(M), Nucleocapsid (N) or Spike (S) proteins. Strong Binder peptides were selected using a %Rank cut-off of 0.5%, which is based on the likelihood of this peptide being presented when compared to a pool of natural ligands. HLA binding predictions performed by netMHCpan were crossed with TAP and proteasome predictions performed by netCTLpan in order to get a more reliable set of peptides. Importantly, the same peptide can be predicted to be presented by more than one HLA.

### Distinct patterns of estimated antigen coverage among populations are observed within different ORFs

The use of predicted HLA-restricted epitopes capable of binding with substantial affinity to the HLA alleles provides a means of addressing population coverage related to HLA polymorphism and different HLA binding specificities. We, therefore, calculated the percentage of individuals from each population predicted to respond to class I SARS-CoV-2 derived SB epitope set on the basis of HLA genotypic frequencies and HLA binding data, using the population coverage calculation tool available through the IEDB. This allowed us to obtain the number of Epitopes/HLA combinations that cover each population at a given rate (Table S3, Supplementary File 3).

We observed an elevated cumulative coverage throughout the countries, spanning from 466.29 in KOR to 565.58 in OMA, with an average of 527.61±24.26, when considering the minimum number of epitopes/HLA combinations predicted to cover 90% of each population. Despite that, significant differences were spotted when evaluating the Area Under the Curves - AUC for distinct populations. The AUC reflects the population coverage by accounting the percentage of individuals capable of presenting an epitope and the number of epitopes/HLA combinations for a given population. By evaluating the estimated antigen coverage we observed distinct patterns across proteins and divergent coverage when comparing distinct populations within proteins (Figure 4, Supplementary File 3). Some populations were better covered for specific portions of the virus, e.g., BRA (AUC=1,438.20) and ITA (AUC=1,408.04) show a high AUC for the protein N (Figure 4D) while others show higher coverages for both N and S proteins, e.g., RSA (AUC=1,614.47 for N, AUC=6,248.83 for S) and ISR (AUC=1431.51 for N, AUC=6,416.80 for S) (Figure 4D, 4F). In contrast, some populations show a high coverage for N protein, e.g. IRL (AUC=1,428.40) and a lower (AUC=5,392.90) for S protein (Figure 4F). Importantly, the minimum and maximum AUC values differ according to each protein as they have a different number of Epitope/HLA combinations (Figure S6).

**Figure 4.**
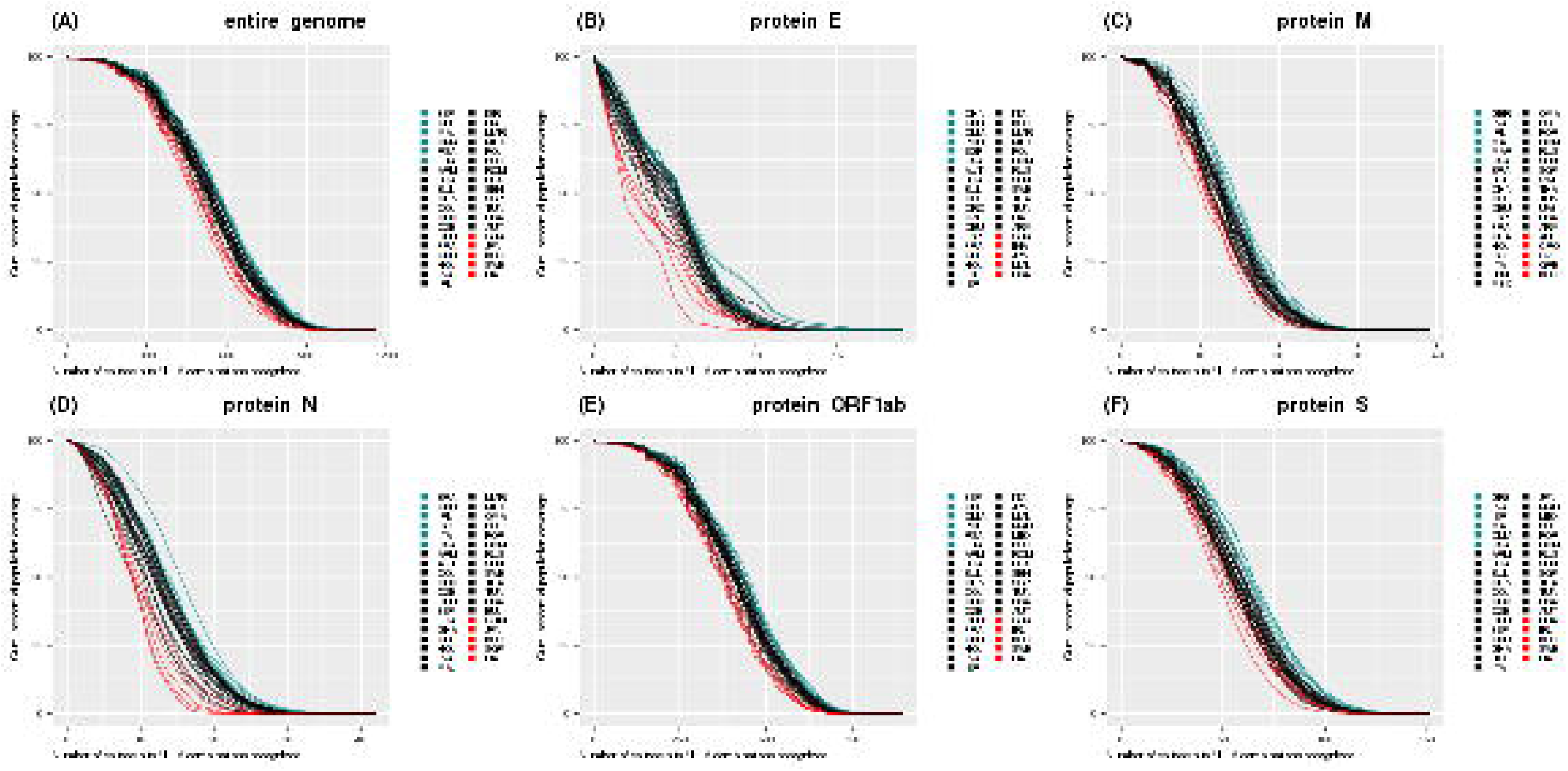
Population coverage for SB peptides derived from SARS-CoV-2 according to the HLA-I set of this study. **(A)** Cumulative population coverage considering the number of possible SARS-CoV-2 derived epitope-HLA allele combinations as a function of the fraction of each population (%). **(B-F)** Same as in **(A)** but for SB peptides derived from specific viral proteins. Each line represents a population and they were colored according to their AUC: the top six higher AUCs are in light blue, the top six lower are in red, and the remaining curves are black. The abbreviations are alphabetically ordered in the legends, not sorted by AUC. E, Envelope; M, Membrane; N, Nucleocapsid; S, Spike protein.

The antigenic coverage within populations can vary since a population is composed of individuals with different ethnic backgrounds. To overcome this limitation, we selected HLA allele from representative studies such as cord blood banks instead of anthropological ones, to investigate antigenic coverage differences regardless of geographical localization, but rather associated with distinct ethnical groups in a given population. Therefore, we performed a population coverage analysis using the HLA alleles described for the four major ethnic backgrounds of the USA: Caucasian, African American, Asian, and Hispanic (Supplementary File 4). The estimated coverage varies among ethnic backgrounds depending on the virus protein analyzed (Figure S7) in which the African American background has less coverage for E, S, M, and ORF1ab proteins, and high coverage for N protein.

### Epidemiological parameters are associated with antigen coverage for specific ORFs

Aiming to understand the relationship between estimated population antigen coverage and COVID-19 manifestations in different populations, we explored associations between the AUC of each country within different ORFs and epidemiological data. We performed Spearman correlations between the epidemiological parameters associated with the COVID-19 pandemic and the AUC of the 5 proteins analyzed and the entire SARS-CoV-2 genome. As explained before, we have chosen the number of deaths per million inhabitants as the main metric (Figure 5), since we believe it may be a more consistent measure, because they are less likely to vary with testing capacity or political strategy. We observed a strong positive correlation between the calculated coverage of each population (represented by the AUCs) and the number of deaths per million inhabitants considering a set with all potential epitopes derived from the SARS-CoV-2 for E (p=0.014, R=0.4, CI=0.0423 to 0.6995) and N (p=0.0028, R=0.48, CI=0.1518 to 0.7036) proteins. Consistently, other significant correlations were also found when considering different epidemiological parameters that are depicted in Figures S8-11.

**Figure 5.**
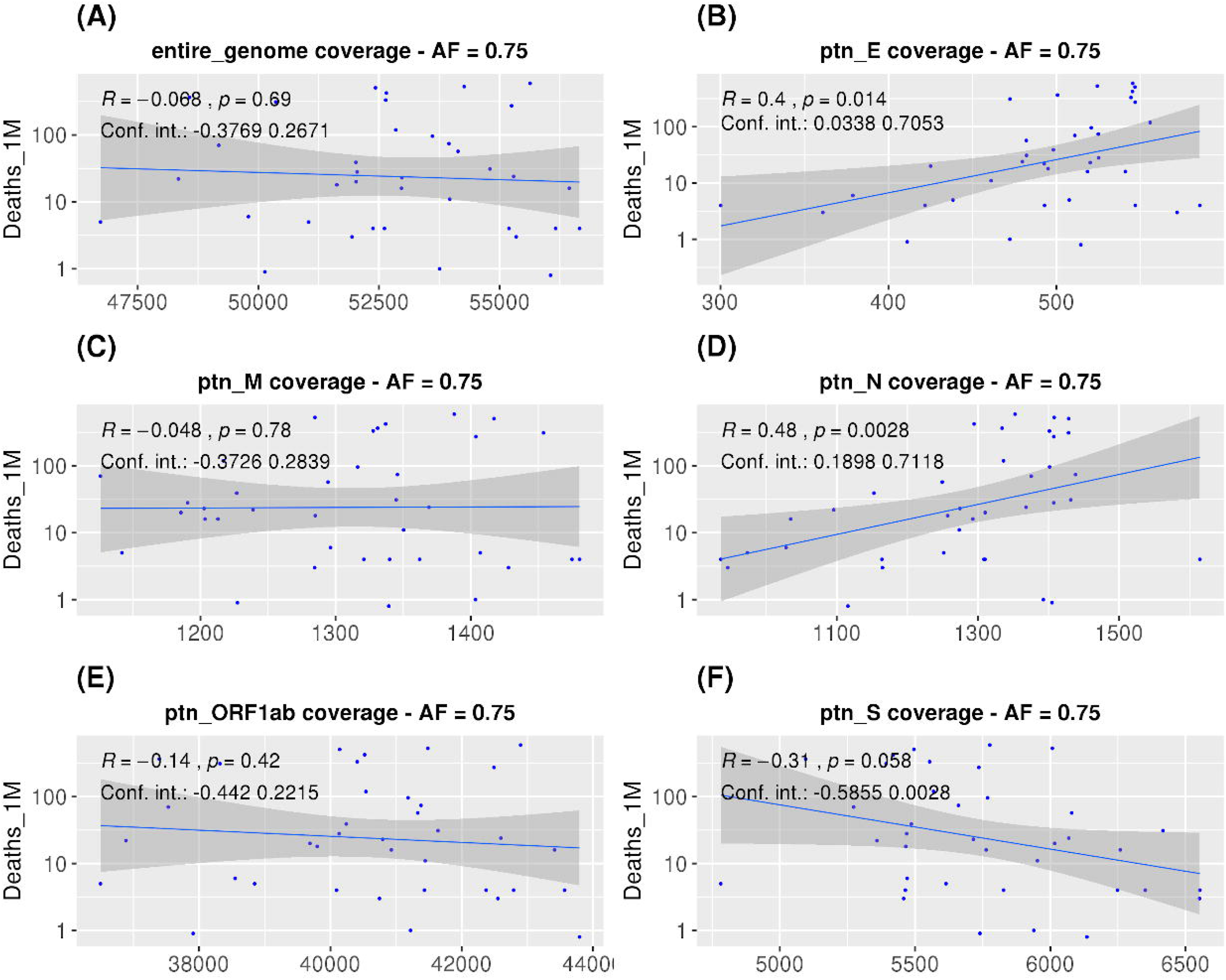
Correlations between SARS-CoV-2 derived SB peptides coverage of each population and COVID-19 outcome. Spearman correlations between AUC calculated from the population coverage considering the entire SARS-CoV-2 genome **(A)**, Envelope **(B)**, Membrane **(C)**, Nucleocapsid **(D)**, ORF1ab **(E)** or Spike protein **(F)** derived peptides and deaths per million inhabitants. For this analysis, a threshold of 0.75 for the allele frequency was considered. The confidence interval shown was calculated using bootstrap with 1000 replacements.

Since we observed inverse correlations for S (borderline p-value) and N coverage associated with the number of deaths per million inhabitants, and because we have observed high HLAs allele frequencies predicted to mainly present S derived peptides in countries from the first quartile, whereas HLAs that potentially present a high number of N derived peptides were enriched in the fourth quartile, we sought to investigate whether distinct patterns of HLA restriction for S and N-derived peptides could better be associated with deaths. In addition, a given population may have an elevated AUC coverage for both S and N proteins, only for S or only for N proteins, and with the ratio, we evaluate the spectrum of S and N derived peptides that simultaneously bind to a collection of HLA alleles. For this reason, we investigated if the ratio between the coverage for S/N proteins is more associated with the number of deaths rather than the coverage of each protein separately. In fact, a stronger association with disease outcome was found when evaluating the ratio between the population’s AUC predicted for S and N proteins together versus the number of deaths per million inhabitants (R=−0.62, p=4.4e-5, CI=−0.89 to −0.3373, Spearman correlation, Figure 6). Of note, the AUC ratio for S and N estimated coverage was 3.84 for African Americans, 4.08 for Caucasians, 4.55 for Hispanics, and 5.15 for Asians. These AUC ratios fall within the limits of previous analysis (3.78 to 5.84, Figure 6) and, although they may indicate a higher number of deaths for people of African American background, the lack of stratified epidemiological data hampered further analysis correlating the ethical group’s coverage with deaths. Therefore, more studies are necessary to confirm these results. Overall, these results highlight the opposite roles associated with potential recognition and presentation of peptides derived from these two SARS-CoV-2 proteins and the COVID-19’s outcome.

**Figure 6.**
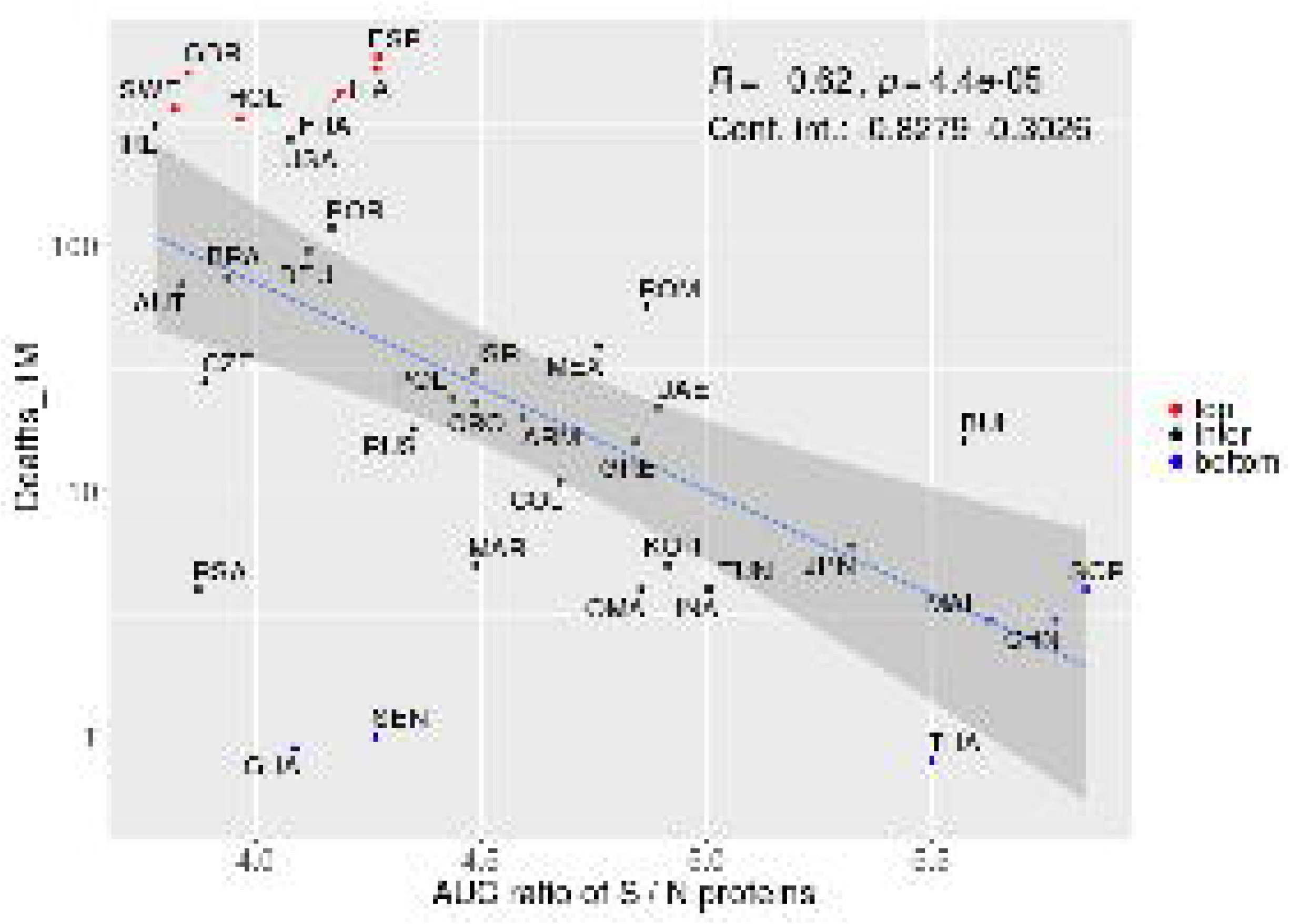
Proteins S and N potentially associated with opposite responses associated with COVID-19. Spearman correlation between the S/N AUC ratio and deaths per million inhabitants. Colors represent countries with a higher (red) or lower (blue) number of deaths. The confidence interval shown was calculated using bootstrap with 1000 replacements.

Interested in better exploring this recognition pattern, we ranked HLA alleles based on the frequency of SB peptides derived from S or N proteins. First, we divided the number of potential binders individually from each protein by their respective lengths as in Table S2 to account for the difference in protein sizes, since bigger proteins tend to generate greater numbers of peptides. Next, for each allele, the normalized frequency of SB peptides in the S protein was divided by the respective normalized frequency in N protein. The majority of HLA-A and B alleles have a ratio superior to 1 meaning that they tend to bind to more SB peptides derived from S than to N derived protein (Figure S12). The alleles with the highest ratios are HLA-A*24:02 (9.54), HLA-A*23:01 (8.87), and HLA-B*18:01 (6.25) in contrast to HLA-A*30:01 (0.49) and HLA-B*07:02 (0.32), showing the lowest ratios. HLA-A*24:02 has an elevated AF in JPN (0.3790) and in many other countries belonging to the first (Q1) and second quartiles (Q2) of deaths per million (Figure 1). On the other hand, low ratio HLA alleles have a higher AF in GHA (0.1107 for HLA-A30:01) and in SWE (0.186 for HLA-B07:02), the latter possessing a higher number of deaths per million than the average. Moreover, HLA-B*07:02 which has the lowest ratio for HLA-B alleles has an elevated AF in several countries of the 4th quartile (Figure S12, Figure 1)

Until this point, we have focused on representing the majority of each population by using an AF of 0.75 that is estimated to represent about 93,75% of the individuals in a given population (PF = 1-(1-AF)^2^) (22). Interestingly, when elevating the HLA allele frequency threshold to 0.9, which is estimated to represent 99% of the individuals, and reperforming the analysis with the same parameters as described for 0.75, a different set of events is observed (Figure S13). To reach the 0.9 threshold, 43 HLA-A and 97 HLA-B alleles were included in which a minimum of 7 HLA-A was used for AUT, JPN and SWE, and 10 HLA-B for SWE, and a maximum of 21 HLA-A and 41 HLA-B alleles were used, both in BRA. The median allele frequency for HLA-A was 0.8997 (0.888-0.910) and 0.8949 (0.8820-0.900) for HLA-B (Table S1). For this analysis, two countries (TUN and THA) that haven’t reached the cutoff of +/- 0.02 deviation for the 0.90 threshold were excluded. The correlations between AUC population coverage and deaths per million inhabitants were lost except for E protein (Figure S13B). Despite the absence of significant correlations between deaths per million inhabitants and AUC coverage for S or N protein, the inverse correlation between AUC ratio for S/N and the number of deaths per million inhabitants (R=−0.43, p=0.01, CI=−0.7587 to −0.0231) is maintained when considering this new threshold (Figure S13G).

To understand the influence of the minor alleles included in the new dataset generated when considering the 0.90 threshold, we performed a new population coverage analysis considering only alleles that were added for this last threshold and are absent from the previous analysis using a threshold of 0.75. Surprisingly, a significant positive Spearman correlation between AUC coverage and deaths per million inhabitants was observed for the S protein (R=0.36, p=0.036, CI=0.0327 to 0.5965) and a negative correlation (R=−0.36, p=0.033, CI=−0.6678 to - 0.0105) for the E protein (Figure S14). Also in contrast to previous findings, a significant positive correlation (R=0.61, p=0.00011, CI=0.3509 to 0.7843) was observed between the AUC ratio for S/N and deaths per million inhabitants (Figure S14G) despite no evident correlation with N protein (Figure S14D).

### Potential peptides for vaccine development

We highlighted the SARS-CoV-2 derived SB peptides generated within each ORF (left Y-axis, Figure 7A) or protein, as well as the number of HLA-I alleles potentially presenting them (right Y-axis, Figure 7A). We observed regions capable of generating an elevated number of unique SB peptides (black bars - top) in comparison to others and the ability of each of these peptides to bind to several HLA-I alleles (blue dots - bottom) which can help to select vaccine candidates. We found 24 unique SB peptides falling within the receptor-binding motif and 3 SB peptides derived from the 4 amino acid insertion (23) in the S1/S2 cleavage site (highlighted in purple in the Figure 7B). These 3 SB peptides vary in the range of affinities and also the number of HLA binders (Table 1). It is also important to note that a 38 amino acid region at the end of S protein (FVSGNCDVVIGIVNNTVYDPLQPELDSFKEELDKYFKN) does not generate any potential HLA-I A or B restricted peptide considering the 140 HLAs analyzed.

**Figure 7.**
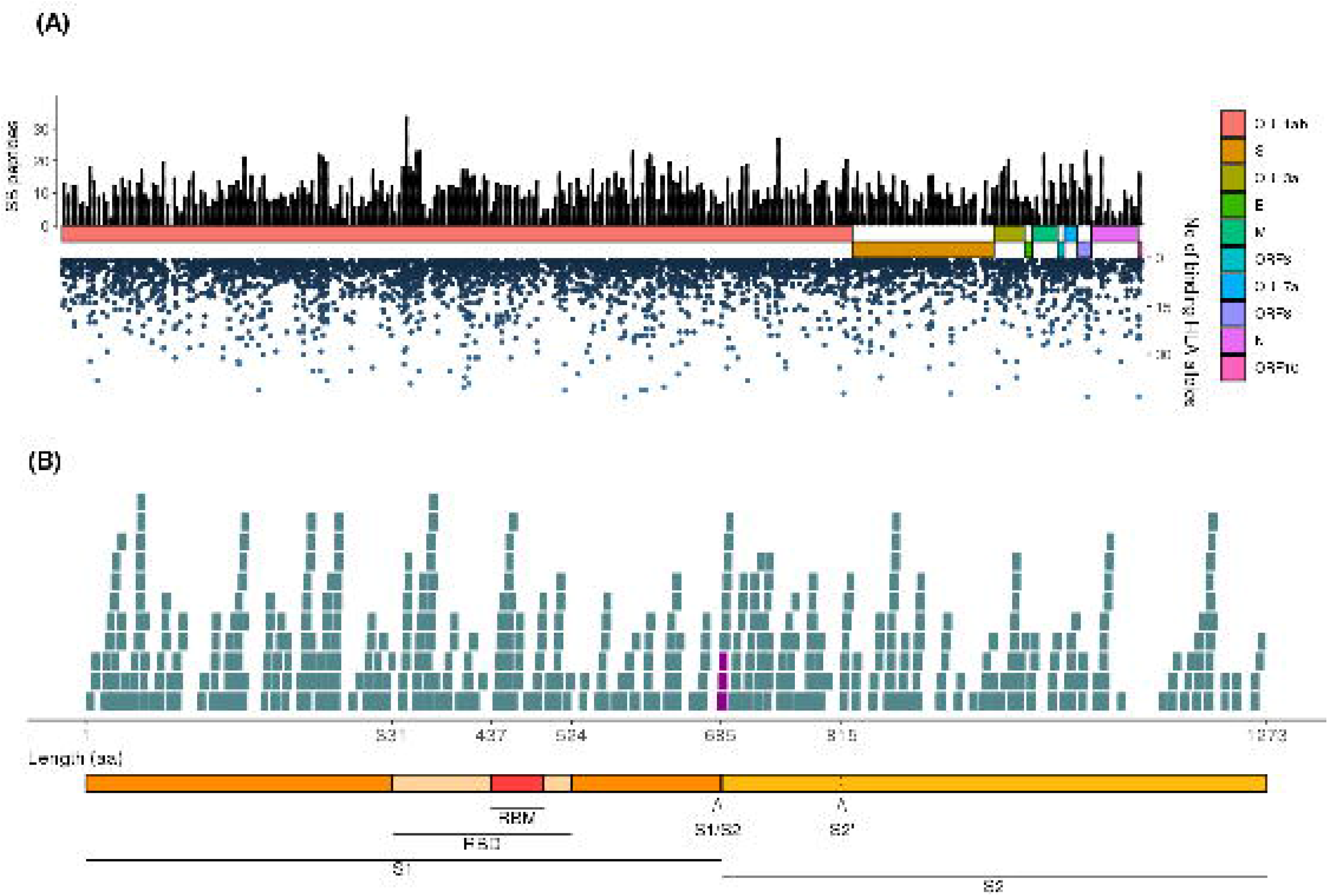
Landscape of SARS-CoV-2-derived SB peptides considering the AF threshold of 0.90. **(A)** The viral proteins and ORFs are represented by colored rectangles proportional to their size. Black bars represent unique SB peptides potentially presented by populations. Blue dots represent the number of HLA-I alleles that a given SB peptide binds to. **(B)** Detail of the S protein and its domains and the SB peptides derived from each region. The seven SB peptides that contain at least one of the 4-length amino acid insertion are highlighted in purple. Length is represented in aa. RBM, Receptor Binding Motif; RBD, Receptor Binding Domain; S1/S2, S1/S2 cleavage site; S2’, S2 cleavage site.

**Table 1.**
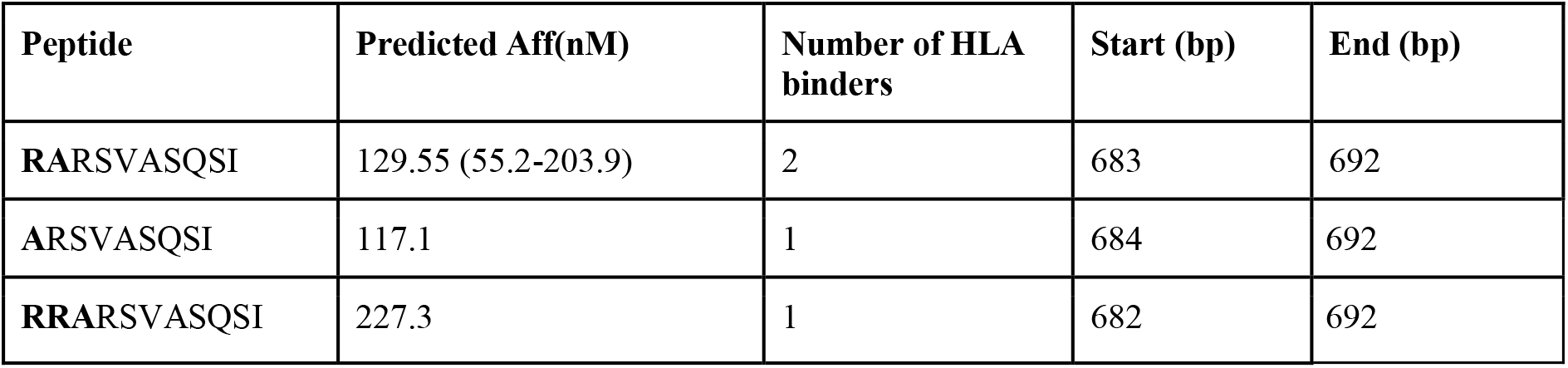
Strong binder peptides including some of the 4 amino acid insertion in protein S, which are highlighted in bold.

| Peptide | Predicted Aff(nM) | Number of HLA binders | Start (bp) | End (bp) |
| --- | --- | --- | --- | --- |
| **R**ARSVASQSI | 129.55 (55.2-203.9) | 2 | 683 | 692 |
| ARSVASQSI | 117.1 | 1 | 684 | 692 |
| **RR**ARSVASQSI | 227.3 | 1 | 682 | 692 |

Aiming to identify candidate peptides to vaccine development, we listed peptides potentially presented by HLA alleles that possess high allele frequencies in countries belonging to Q1 (Figure S15) including but not limited to HLA-A*24:02, HLA-A*23:01, HLA-B*15:02, and HLA-B*58:01 while avoiding alleles prevalent in Q3 and Q4. It is important to mention that, as stated previously, an unfavorable outcome was observed for countries with high AF for HLA-A*02:01. Therefore, aiming a vaccine design based on peptides predicted to be presented by this allele may prove ineffective in some populations. A total of 126 candidate peptides were identified considering 85 HLA alleles (Figure S15, Supplementary File 5). Six peptides were selected to estimate the global populational coverage considering all deposited populations on IEDB. A coverage of 21.09% is achieved with one peptide, 42.78% with two peptides, and up to 79.99% with 6 peptides (Table 2).

**Table 2.** Global population coverage analysis based on the allele frequencies available at the Immune Epitope Database.

| Peptides | Projected population coverage | Average number of epitope hits / HLA combinations recognized by the population | Minimum number of epitope hits / HLA combinations recognized by 90% of the population |
| --- | --- | --- | --- |
| TRFQTLLAL | 21.09% | 0.22 | 0.13 |
| TRFQTLLAL<br>YFPLQSYGF | 42.78% | 0.51 | 0.17 |
| TRFQTLLAL<br>YFPLQSYGF<br>SVLNDILSR | 54.12% | 0.68 | 0.22 |
| TRFQTLLAL<br>YFPLQSYGF<br>SVLNDILSR<br>ATRFASVYAW | 64.66% | 0.9 | 0.28 |
| TRFQTLLAL<br>YFPLQSYGF<br>SVLNDILSR<br>ATRFASVYAW<br>KEIDRLNEV | 75.51% | 1.18 | 0.41 |
| TRFQTLLAL<br>YFPLQSYGF<br>SVLNDILSR<br>ATRFASVYAW<br>KEIDRLNEV<br>ESNKKFLPF | 79.99% | 1.32 | 0.5 |

Altogether, these results indicate a distinct pattern of virus recognition and response among populations. HLA allele profiles and predicted antigen coverage for specific ORFs show a strong correlation with mortality by COVID-19. These results highlight the relevance of distinct responses to proteins S and N among different populations to COVID-19 pandemic evolution.

## Discussion

In this work, we investigated the distinct susceptibility associated with HLA diversity profiles among populations facing the SARS-CoV-2. We applied multiple *in silico* approaches to predict the population’s response to the SARS-CoV-2 infection and correlated these findings with the number of deaths per million and other epidemiological parameters. We suggest that a positive country outcome relies partially on the ability of the majority of a population to present peptides derived from the S protein whilst avoiding those derived from the N protein, as it correlates with an unfavorable outcome.

Numerous factors such as political rules implementation, adherence to social distancing measures, health system structures, and population density can affect the spread of SARS-CoV-2 and impact COVID-19 outcome. Besides, underreporting and testing policies vary among countries and also influence the epidemiological reports (i.e.: INA, MEX, SEN, JPN, MAR TUN, BRA, COL, and THA performed tests in less than 1% of the population). Absolute epidemiological parameters such as total number of cases, serious/critical cases, deaths or recoveries are biased by the number of cases or population size. In this sense, when using the number of deaths per number of cases, the countries’ testing policy can directly interfere with the data because countries with an open testing policy tend to have fewer serious cases and deaths than countries which test only hospitalized or risk group individuals. Therefore, we choose to use deaths per million inhabitants as the main metric since it also has been used as a proxy for disease outcome (24,25). In addition, other factors like blood type and ethnicity (26,27), countries BCG vaccination policy (24), or the percentage of obese people (28) could influence mortality, affecting the relationship between deaths per million and Spike or Nucleocapsid proteins AUC coverage. In fact, some countries (e.g.: ITA, ESP, FRA, GBR, USA, and HOL) showed higher deaths per million than expected by the relation between AUC ratio of S/N and deaths per million inhabitants. Considering that, we included as many countries as possible in the analysis to minimize these discrepancies rather than normalize for the particularities of each population.

Our analyses depict the potential population’s patterns of SARS-CoV-2 recognition of class I HLA restricted peptides. The HLA alleles repository allowed us to carefully select populations prioritizing blood donor registry rather than anthropology studies to better represent a country. Since the most relevant genes for CTL responses are the HLA-A and B and considering that HLA-C data is not available for several populations, we have not accounted for HLA-C (29). It also allowed us to characterize in detail the potential responses associated with major and minor alleles. Remarkably, HLA-B*15:03, the allele predicted to present more peptides in previous works (12,13), ranks 12th out of 76 HLA-B alleles analyzed. As the methodology and metrics used in these previous works differ from ours, different results are expected.

HLA systems have a strong linkage disequilibrium (LD) that can result in conserved multi-locus haplotypes, mostly because the alleles are arranged in adjacent loci in a specific region of the chromosome, where the distance between them, among other factors, can affect the linkage disequilibrium (30), and, consequently, their random inheritance in a given population. To correct for LD, we would need to have access to full haplotypes frequencies instead of allele frequencies, which is not available or incomplete for many countries impacting the size of the study. Despite that, although our study does not consider the LD of the HLA system, we believe that using high AF thresholds, as we did, minimize the need for the correction for LD. Furthermore, the impact of LD on the calculation of population coverage by IEDB has been considered negligible (14).

HLA binding predictions have become a useful and important method for selecting promising peptides for vaccination. Moreover, by comparing the predicted binding affinities with the affinities verified through assays deposited at the IEDB, we showed that SB peptides, the focus of this study, have comparable affinities to the peptides experimentally defined as high and intermediate binders. In addition, their binding affinities are in the range of those peptides known to trigger a TCR response as detected by T cell-based assays as opposed to WB peptides. Previous works have highlighted the importance of population coverage studies to uncover candidate peptides for vaccination. The inverse correlation between coverage for proteins S and N and deaths per million inhabitants corroborates a study showing that CD8 T lymphocytes from convalescent patients display more reactive CD8+ T cells for S than for the N protein (10), highlighting its immunogenic role. This same report indicates that several immunogenic HLA-I restricted peptides are derived from S protein, in line with our findings that S peptide coverage has a positive impact on COVID-19 outcome. These results suggest CD8 mediated anti S-derived peptide responses could be one of the mechanisms associated with more favorable COVID-19 outcomes.

Features of SARS-CoV-2 S proteins are responsible for unique characteristics of this new virus. A recent report indicated differences in the protein S sequence regarding the receptor-binding motif and a four amino acid insertion when comparing with bat and previous strains (23). We found unique SB peptides falling within the receptor-binding motif sequence, including the 4 amino acid insertion in the S1/S2 cleavage site. This sequence is considered to be related to the high virulence of SARS-CoV-2 (23) and HLA-I restricted peptides derived from this region could help mounting differential immune responses between SARS-CoV-2 and other coronaviruses. On the other hand, the lack of potential peptides derived from the FVSGNCDVVIGIVNNTVYDPLQPELDSFKEELDKYFKN region is somehow intriguing and exploiting functional features of this segment and evolutive forces that could have shaped its low immunogenicity can shed light over the relevance of these findings.

The scarceness of N protein derived immunogenic HLA-I restricted peptides reported by Grifoni and colleagues (2020) resembles the low proportion of N protein-derived peptides found in our analysis. This reinforces the concept that N derived peptides may not be involved in functional and productive anti-SARS-CoV-2 responses since our correlation analyses indicate that populations with greater N coverage have higher numbers of deaths per million inhabitants. Protein N is highly expressed and has a high immunogenic potential so that antibodies against it are the first found in the serum of infected patients (31). Since the homology between N proteins of SARS-CoV-2 and SARS-CoV is over 90% (15) and the SARS-CoV-2 peak viral load is earlier (32), the immunogenicity of SARS-CoV-2 protein N is believed to be similar to that of SARS-CoV with an even faster response. However, when investigating T lymphocytes from convalescent patients, a lower frequency of targets associated with protein N was observed (10), indicating that these peptides are either less immunogenic or could lead to the depletion of reacting cells by some T cell hyperactivation mechanism. If this is the case, N derived peptide-based hyperactivation could be related to cytokine storm, a process largely associated with the most severe cases of COVID-19 (4,33,34). The number of deaths per million inhabitants showed a significant positive correlation when correlated with N coverage, reinforcing the possibility that N peptides lead to a more aggressive disease. This dysregulation of the immune system has been shown to occur in other viral infections and could be dependent on CD8 T lymphocytes (35).

Assuming that, like rhesus monkeys, individuals previously infected with SARS-CoV-2 are protected from reinfection, herd immunity becomes possible for COVID-19 (36). Herd immunity consists of individuals being protected from infection by being surrounded by immune non-infected individuals, therefore unable to transmit the disease (37). Previous studies have shown that the average number of SARS-CoV-2 transmission caused by a single infectious individual (R_0_) is from 2 to 6 (38,39), so assuming a R_0_ of 4, the herd immunity would be achieved with 75% immune individuals. By using a threshold of 0.75 for cumulative allele frequencies for HLA-A and B within each population, we estimate that 93.75% of the country’s population is covered by our analysis. In this context, vaccine strategies based on S derived peptides would induce protective responses to the infection in the majority of the population that carry more frequent HLAs. The opposite would be expected regarding the minority, carrying rare HLAs, that display a positive correlation between S coverage and the number of deaths. On the other hand, the same rationale may apply to N and E proteins which correlate positively with the number of deaths for the major HLA alleles but display an inverse tendency when only the minor alleles are accounted for. In summary, an elevated S/N coverage for the most frequent HLA alleles showed protective effects whilst lower S/N coverage ratios were protective for the minor HLA alleles within populations. It is important to mention, though, that reaching herd immunity through vaccination for COVID-19 can be a hard task, probably requiring revaccination, since the immune response to SARS-CoV-2 may be lost rapidly (40) and people might thereafter be susceptible to reinfection with the virus.

A country’s population is normally composed of multi-ethnic groups and backgrounds, which could undergo distinct susceptibilities to SARS-CoV-2 infection. In addition, there is an increasing debate on whether some ethnic groups are being disproportionately affected by COVID-19, which is frequently enhanced by distinct access to the health system and social inequities (27,41,42). Our attempt to investigate antigenic coverage within ethnic groups of the same country aimed to check differences in their coverage patterns, regardless of geographical localization. Overall, our results suggest that differences within ethnic backgrounds may exist and could be associated with a better or worse susceptibility for these groups. However, more studies are necessary to confirm this pattern, and also to explore these discrepancies in other countries since the ethnic background should differ from country to country. The results also highlight the need for distinct political strategies to deal with the pandemic, which differs among countries. We were able to identify two HLA alleles that are highly prevalent in countries showing less number of deaths and may have protective effects regarding the immune response to SARS-CoV-2. The vast majority of peptides derived from protein S potentially presented by those two alleles were absent from supertype HLAs such as HLA-A*01:01, A*02:01, A*03:01, B*15:01, and others. In fact, the most prominent worldwide allele HLA-A*02:01 potentially presents peptides from the ORF1ab region. A recent study has shown that this region is not immunodominant in CD8-mediated immune response when challenged with SARS-CoV-2 Epitope MegaPool in convalescent donors (10). HLA-A*24:02 is highly frequent in many populations and the predictions made for this allele could help drive vaccine development.

Finally, we highlighted some important viral regions predicted to be presented by a vast number of HLA alleles which can help to understand the response to viral peptides and peptide vaccine design. More importantly, we suggest that peptide-based vaccination strategies should rely mainly on the S protein due to its negative correlation with the number of deaths per million inhabitants and its role in the infection. On the other hand, we present a cautionary note for using the N protein since it showed a strong positive correlation with the number of deaths per million when considering individuals carrying frequent alleles. Last, we identified potential antigens derived from the 4 amino acid insertion of SARS-CoV-2 that is absent from previous strains and could be used for studies based on T cell response, as it would allow differentiation from previous coronavirus infections and importantly, serve as a guide for SARS-CoV-2 specific vaccine development. To our knowledge, this is the first work to focus on antigen population coverage associating with COVID-19 outcome.

## Material and Methods

### Epidemiological data

The epidemiological data used in this analysis (number of total cases, total deaths, total recovered, serious critical cases, total cases per 1 million of inhabitants and total deaths per 1 million inhabitants) were retrieved on the Worldometer database (www.worldometers.info/coronavirus, accessed at 01:18PM, May 17, 2020). This database collects data from official reports, directly from Government’s communication channels or indirectly, through local media sources when deemed reliable. The 50 countries with the highest death per million inhabitants and more than 1 thousand total cases were evaluated.

### HLA-I gathering

Human Leukocyte Antigen (HLA) I alleles for A and B genes with at least two fields and its frequencies in the selected populations were retrieved from the Allele Frequency Net Database (43) between 23/04/2020 and 07/05/2020. Larger datasets that were not part of anthropological studies nor minority ethnic groups, and with at least 50 individuals were preferred (Supplementary File 2). The alleles were ordered from the most frequent to the least frequent and the allele frequencies were summed to obtain a cumulative allele frequency of 0.75 or 0.90 separately for HLA-A and B. The values used were the closest to the threshold (equal, above or below) considering the difference between the cumulative allele frequency and the threshold itself. Countries that haven’t reached these thresholds with a difference inferior to 0.04 for AF=0.75 and 0.02 for AF=0.9 were excluded. The minor alleles (AF=0.15) are the alleles exclusive from the dataset generated when using 0.90 as a threshold. Allele frequencies with resolution greater than two fields were combined, e.g. for BRA: B*07:02:01 and B*07:02:03 became B*07:02. The HLA alleles for 4 ethnic backgrounds (African American, Caucasian, Hispanic, and Asian) were obtained for the USA population and analyzed as previously described.

### Accessing experimental binding affinities

We searched at the Immune Epitope Database (IEDB) on 18/05/2020 for MHC Ligand assays that matched the parameters “Linear Epitope”, Organism “Virus”, positive assays only, MHC Ligand Assays, MHC Restriction: “MHC Class I”, HLA-A and HLA-B, Host: “Humans”. Assays with NA values for “Quantitative.measurement” and entries with “>”, “≤” or “<” in "Measurement.Inequality" were excluded. Also, we retrieved T cell assays using the same parameters except for the positive assays only.

### Binding predictions

The translated proteome from SARS-CoV-2 genome version BetaCoV_Wuhan_IPBCAMS-WH-01_2019 was downloaded on 23/03/2020 from the NCBI repository (https://www.ncbi.nlm.nih.gov/nuccore/MT019529). Binding predictions were performed using netMHCpan4 (19) with peptide lengths from 8 to 11 for the entire viral genome. We used blastp v2.9.0 to eliminate peptides that matched protein sequences from the Ensembl human genome GRCh38 with 100% of identity and same query length. Next, we performed predictions with netCTLpan to also remove peptides with a TAP score smaller than 0 and a proteasome cleavage score smaller than 0.5 to assure that our analysis only includes peptides with a high probability of being processed by proteasome and TAP. Only strong binder peptides (%Rank <0.5) were kept for further analysis.

### Population Coverage

We used the tool described by Bui and collaborators (14) available at IEDB (http://tools.iedb.org/population/) to calculate the population coverages. The inputs used were the same allele frequencies for the binding predictions (Table/Supp file) and the Strong Binder peptides for each HLA-I allele. The analysis was performed using the “Class I separate” option. Coverages were calculated for the entire SARS-CoV-2, and separately for S, N, M, E, and ORF1ab viral proteins. Area Under the Curve (AUC) was calculated prior to Spearman correlations.

### Statistical analysis

Significant results for correlations were considered when p-value < 0.05. A bootstrap with 1000 replicates was performed to access the confidence interval for Spearman correlations with epidemiological data. We used the complete method and 1-Pearson correlation coefficient as the distance to cluster rows and columns of the heatmaps. Cluster stability for heatmaps was calculated using Jaccard distancing through bootstrapping using 10,000 replacements and cluster method “disthclustCBI”. Spearman correlations were used unless stated otherwise.

### Data analysis

The analysis was conducted using the R environment version 4.0 and the following packages ggpubr v0.2.1, ggplot2 v3.2.1, data.table v1.12.2, gridExtra v2.3, readxl v1.3.1, dplyr v0.8.3, tidytext v0.2.3, pheatmap v1.0.12, RColorBrewer v1.1, cowplot v1.0, GenomicRanges v1.31.1, IRanges v2.18.1, stats v4.0.2, boot v1.3, ggfortify v0.10 and ggbio v1.32.

## Data Availability

Data will be available after final publication.

## Acknowledgments

MAMP was supported by CAPES fellowship program. The authors also would like to thank the Bioinformatics Core Facility (INCA-RJ) for their support.

